# Changes in Walking Energy Expenditure and Substrate Oxidation during Pregnancy

**DOI:** 10.64898/2026.06.28.26356764

**Authors:** A.M. Cagiao, J. Fariñas, J. Rial-Vázquez, M. Rúa-Alonso, M. A. Giráldez-García, A. M. Jácome, M. L. Erickson, E. A. Carnero

## Abstract

**Introduction:** obesity and unhealthy weight gain during pregnancy are associated with risk of pregnancy complications. Management of energy balance during pregnancy needs an accurate assessment of intake and energy expenditure. As pregnancy promotes specific physiological changes, energy expenditure during rest and activity throughout this period may be altered. The aim of this study was to estimate with accurate methods changes in energy expenditure, substrate oxidation and efficiency during the most common activity in pregnancy that is walking.

**Methodology:** it was a prospective observational study during pregnancy. A graded steady state walking submaximal exercise test was used to calculate indirect calorimetry variables with a portable metabolic cart at each trimester of pregnancy in 30 healthy pregnant women. Walking energy expenditure (WEE), carbohydrate (CarbOx) and fat oxidation (FatOx) were calculated with exercise intensity specific stoichiometric equations from measured oxygen consumption (VO₂) and carbon dioxide production (VCO₂). Resting component was removed from WEE to calculate net walking energy expenditure (Net WEE). Moreover, participants were classified as having healthy or unhealthy gestational weight gain (GWG) according to the Institute of Medicine (IOM) recommendations. Changes in body weight during the study were used to adjust net WEE and substrate oxidation. Walking exercise efficiency was calculated as work rate (WR) divided by WEE or net WEE. Differences in main variables during pregnancy were analyzed using a general lineal model. Least square means analyses were utilized to compare differences in WEE, Net WEE, and substrate oxidation between healthy and unhealthy weight gain groups.

**Results:** Net WEE increased significantly during pregnancy. Higher rates were found between the second and the third trimester of pregnancy at any given speed. This elevation was reflected in increased FatOx during moderate intensity exercise and higher CarbOx at the fastest walking speed. Therefore, a significant interaction time*speed for NetWEE and substrate oxidation was found. Weight gain was an important variable in energy expenditure and substrate oxidation as quantitative differences in NetWEE and CarbOx were due to change in weight registered during the study. Changes in Net WEE throughout pregnancy were higher in the unhealthy GWG group which expended significantly more CarbOx than the healthy group at each speed. Overall, efficiency decreases during pregnancy (1.48% from the second to the third trimester and 1.88% from the first to the third one) and pregnant women were more efficient in moderate intensity exercise (walking at 4 km/h) independently of the trimester of pregnancy.

**Conclusion:** The results suggest a paradoxical intensity-dependent substrate oxidation selection during walking exercise throughout pregnancy as in no-trained pregnant women the increase in WEE relies on fat oxidation in a short bout of activity. The results may be relevant for the management strategies in obesity and excessive GWG during pregnancy.

## Introduction

Obesity in pregnancy has increased in parallel with the increased prevalence of obesity in the general population and is projected to continue to increase (1). Maternal pre-pregnancy body mass index (BMI) and gestational weight gain (GWG) are associated with risks of pregnancy complications. Achieving a healthy pre-pregnancy BMI and GWG may reduce the burden of pregnancy complications and ultimately the risk of maternal and neonatal co-morbidities (2). A combination of diet, exercise, and behavioral modification must be part of the standard care to modify energy balance components (3), which may be also applicable for pregnant women. Therefore, clinical assessment of energy expenditure and energy intake must be a priority to an exhaustive management of energy balance during pregnancy.

Since physical activity is the most variable component of total energy expenditure (4), an accurate assessment of physical activity energy expenditure (PAEE) must be necessary to prescribe energy needs during pregnancy. However, pregnancy promotes specific physiological changes (cardiovascular, respiratory, hormonal, and metabolic adaptations, increase in body mass and modifications of motor patterns), to meet the increased metabolic demands of the mother and fetus which may affect the relationship between exercise energy expenditure and external load (4). Thus, estimations of energy expenditure from an external load may be inaccurate during pregnancy.

Few studies have investigated physical activity energy expenditure in pregnancy with accurate methods (5). Therefore, we decided to study changes in exercise energy expenditure during pregnancy in the most common activity in pregnancy which is walking. We hypothesized that specific physiological changes occurring during the pregnancy will affect walking energy expenditure (WEE), substrate oxidation and walking efficiency (ExE) independently of weight change during pregnancy. Additionally, we believe that excess weight gain during gestation may affect WEE, substrate oxidation, and ExE.

Our main goal was to analyse WEE, substrate oxidation and ExE in healthy pregnant women throughout the three trimesters of gestation. Also, we aimed to compare changes in WEE, substrate oxidation and ExE between women with excessive and normal weight gain during the pregnancy.

## Methods

### Design

This was a prospective follow-up study during pregnancy to analyze changes in WEE, substrate oxidation and ExE. Participants were recruited between April 2021 and September 2022 to perform 3 cardiopulmonary submaximal exercise tests, one at each trimester of their pregnancies. At the screening visit, clinical history and risk factors were evaluated by obstetricians and participants’ midwifes, who authorized them to perform moderate exercise. Women who were taking medication that could alter metabolism, who had diabetes prior to pregnancy, severe anemia, or uncontrolled thyroid disease, as well as those who consumed drugs, alcohol, or tobacco, were excluded. All procedures included in the study were approved by the ethical committee of clinical research in Galicia (Spain) (2019/2013) and an Institutional Review Board of Faculty of Sport Sciences and Physical Activity at the University of A Coruña (Spain). After the screening visit at their health care center, participants signed an informed consent form (ICF) to approve enrollment in the study. Moreover, signed ICF included authorization to have access to their medical records and clinical history for research purposes.

The study consisted of a screening visit, 3 one-day study visits, 3 control visits and a variable number of follow-up health pregnancy visits. All assessments were completed in the same laboratory at University of A Coruña (Spain) before 12, between 16 and 18 and between 32 and 35 gestation weeks (Figure 1). During the screening visit participants were instructed about the protocol they must follow for the study visits. Briefly, participants were asked to eat a light meal 3 hours before the visit and not consume caffein beverages on the assessment day; also, they had to wear comfortable sport shoes and light clothes, to be well hydrated and rested. A PAR-Q questionnaire was completed by each woman in visit 1 to confirm their safety to engage in moderate-vigorous PA and discard medical contraindications to exercise. Transportation was guaranteed by participants’ request.

**Figure 1.**
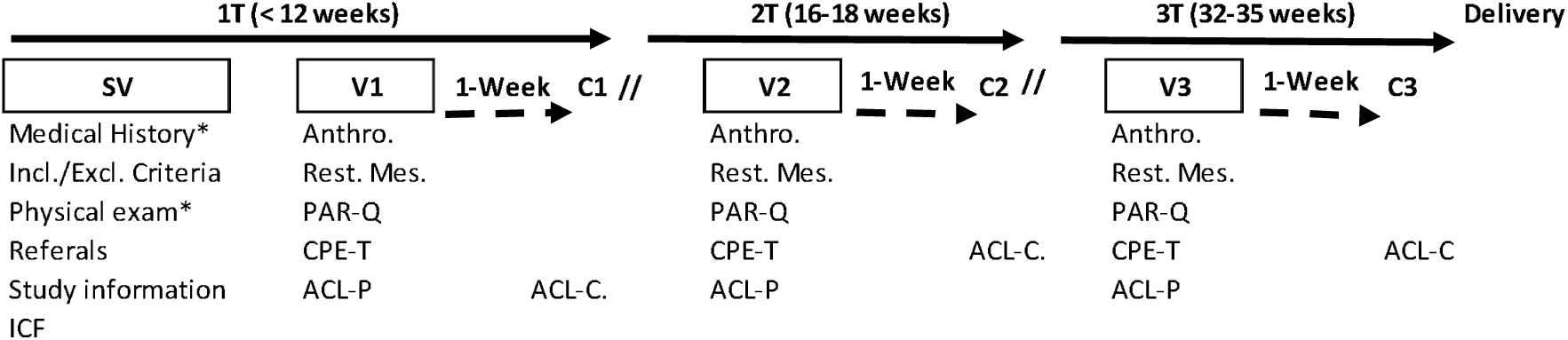
Study timeline. 1T, first trimester; 2T, second trimester; 3T, third trimester; SV, screening visit; V1, visit 1; V2 visit 2; V3, visit 3; C1, control visit 1; C2, control visit 2; C3, control visit 3; Anthro, Anthropometric measurements; Rest. Mes., Resting measurements; PAR-Q, Physical activity readiness questionnaire; CPE-T, Cardiopulmonary exercise test; ACL-P, accelerometer placement; ACL-C, accelerometer control; ICF, informed consent form.

#### Participants

50 healthy pregnant women who visited a health care center before 12 weeks of gestation accepted to participate in the study; however, only 30 completed the full protocol (Supplemental figure 1, Supplemental Digital Content). After the screening visit and during the follow-up visits, was confirmed that all women had low-risk pregnancies with no contraindications to exercise. All participants delivery at term, 25 had spontaneous delivery and 5 underwent cesarean section; All newborns were healthy and did not require additional care or support. During the pregnancy, 3 (10 %) pregnant women had gestational diabetes without insulin treatment needed, 4 (13.3%) had not complicated gestational hypertension and 1 (3.3%) developed fetal growth restriction (this condition was detected after the third assessment).

#### Study visit protocol

The visit began with a brief anamnesis conducted by the study physician to confirm non-acute contraindications to complete the cardiopulmonary exercise test (CPE-T). Afterwards, weight and standing height were measured to the nearest 0.1 kg and 0.1 centimeters, respectively with Añó Sayol Barcelona scale and stadiometer. After a sitting resting period, blood pressure was measured manually in both arms by a trained nurse before each test according to American Heart Association guidelines (6). Lab temperature and humidity were taken before each measurement with a digital thermos hygrometer (TFA Dostmann GmbH &Co. KG,Zum Ottersberg 12, 97877 Wertheim, Germany; visit 1= 21.7 ± 1.8°C and 51.7 ± 9.1%; visit 2= 21.9 ± 1.0 °C and 51.4 ± 9.4%; visit 3= 22.1 ± 2.0°C and 53.9 ± 9.3%).

#### Energy Expenditure and Substrate Oxidation Assessments

A graded steady state walking exercise on a treadmill (Technogym excite med L1, Technogym, United Kingdom) was utilized as mode of exercise with simultaneous measurement of oxygen uptake (VO_2_), CO_2_ output (VCO_2_) and ventilatory variables with a portable metabolic cart (Cortex Metamax 3B) and heart rate (HR, Polar H10 (Kempele, Finlandia)). At the beginning of each CPE-T, environment conditions of the room were measured to confirm participants perform test under thermoneutral conditions. Gas analyzers and flow sensor were calibrated following the manufacturer recommendations with a reference gas tank and 3-L syringe, respectively. Before the gases data collection, a basal EKG was recorded and supervised by the study physician to clear the participant to start the CPE-T. The protocol started with a 5-min resting period (3-min sitting and 2-min standing) followed by 4-min consecutive stages walking on a treadmill at 2 ,4 and 6 km/h (S1, S2, S3 respectively) with 1% slope. The last-minute data of each stage was used to calculate all indirect calorimetry variables. Rated perceived exertion (RPE) was evaluated at the end of each stage by Borg scale (graded 1-10) (7). Participants were not allowed to grasp the handrails while walking. There were 2 minutes of recovery after the exercise was done, walking at 2 km/h.

#### Energy Expenditure and Walking Exercise Efficiency Calculations

WEE, carbohydrate, and fat oxidation rates were calculated with stoichiometric intensity specific equations from measured VO_2_ and VCO_2_ (8). These calculations assume negligible contribution of protein oxidation (see equations 1-6). Additionally, resting energy expenditure component (sitting) was removed from WEE to calculate net walking energy expenditure (Net WEE). Either WEE and Net WEE were expressed in kcal/min and carbohydrates and fat oxidation in mg/min.

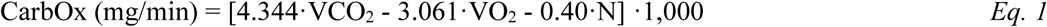

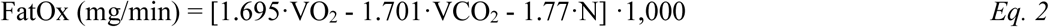

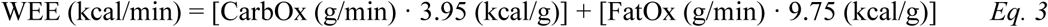

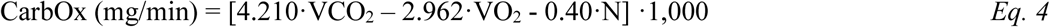

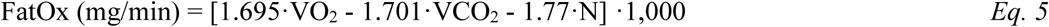

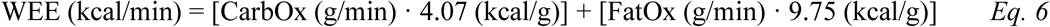

VO_2peak_ was estimated by simple linear regression analysis following the procedure of the American College of Sport Medicine (9). Maximum HR was calculated from Tanaka’s equation (10).

#### Walking Exercise Efficiency

ExE was calculated by dividing work rate (WR, equation 7) over WEE (Gross ExE) or Net WEE (Net ExE). Additionally, we calculated delta efficiency using the slope of the regression between WR and WEE (Delta ExE) . All three exercise efficiency variables were expressed in percentage (11).

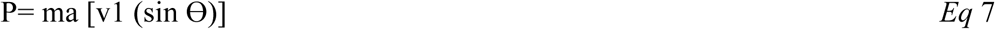

*where m is the mass (kg) of the woman, v1 is the treadmill speed (m/s), a is the gravitational constant (9.8 m/s), and sin Ɵ represents the treadmill grade*.

#### Total Daily Physical Activity Assessment

Participants wore an accelerometer (GT9X Actigraph) on their non dominant wrist for 7 days after each walking test to quantify total daily physical activity (TDPA, steps/day). The accelerometer sampled at 30 Hz, using a normal filter and an epoch length of 60 seconds. To be considered a valid assessment, a minimum of 21 hours of wear time per day and at least three valid weekdays and one valid weekend day of recordings were required.

#### Weight gain variables

We calculated two weight gain variables. Total gestational weight gain (GWG) was defined as the difference between the weight measured at the screening visit and at the last visit before delivery. Participants were stratified into healthy and unhealthy GWG groups according to the Institute of Medicine (IOM) recommendations, based on early pregnancy BMI categories (12). Additionally, the weight difference between visit 1 and visit 3 (ΔWG 3T-1T) was used to adjust for changes in weekly energy expenditure (WEE) and substrate oxidation.

#### Statistical Analysis

Data are summarized as mean and standard deviation (SD) or standard error (SE) always variables were normally distributed. Variables non-normally distributed were transformed into natural logarithm to achieve normal distribution. Differences in WEE, HR, respiratory exchange ratio (RER), substrate oxidation and ExE variables along the three trimesters of pregnancy were analyzed using a general linear model. Additionally, to account for the effect of weight gain in WEE and substrate oxidation, an ANCOVA repeated measures analyses was utilized to compare the differences in WEE and substrate oxidation between healthy and unhealthy weight gain group and by adjusting to ΔWG 3T-1T (in absolute (kg) and relative values (%)).The least square means analyses were obtained for each group, trimester and speed after adjusting for each individual covariable (including all possible interactions); least square means differences between groups were analyzed using Bonferroni’s test.

All statistical procedures were performed using IBM-SPSS® 28.0 (IBM, Chicago, IL, USA) and GraphPad Prism – for Windows® (GraphPad Software Inc.).

## Results

The characteristics of the participants that completed the three assessments are described in table 1. Out of the 30 women evaluated at the beginning of the pregnancy 43.3% were normal weight, 33.3% overweight and 23.3 % obese. BMI increased during pregnancy and ΔWG 3T-1T was above 15% on average, although it was highly variable and 43% of the participants had an unhealthy GWG at the end of the pregnancy (n = 13, these participants were over the recommendations, no woman was below). Resting HR was significantly higher during the third trimester than in the first and second one, which resulted in diminution of the HR reserve at the end of the pregnancy. Estimated absolute VO₂_peak_ increased significantly during the pregnancy (*F*= 4.196, *P=*0.032). Total daily physical activity (steps per day) decreased significantly from the second to the third trimester (table 1).

**Table 1.** Descriptive characteristics of study participants (n=30) throughout the three trimesters of pregnancy.

|  | 1T | 2T | 3T |
| --- | --- | --- | --- |
| Age (years) | 33.2 ± 4.9 |  |  |
| Gestational week (weeks) | 10.5 ± 1.5 | 17.5 ± 1.4 | 32.8 ± 1.6 |
| BMI (Kg/m<sup>2</sup>) | 26.5<sup>a**</sup> ± 5.2 | 27.4<sup>b**</sup> ± 5.3 | 30.3<sup>c**</sup> ± 5.3 |
| HR resting (beats/min) | 81<sup>a**</sup> ± 14 | 81<sup>a**</sup> ± 11 | 92<sup>b**</sup> ± 10 |
| HRR (beats/min) | 104<sup>a**</sup> ± 13 | 104<sup>a**</sup> ± 11 | 93<sup>b**</sup> ± 10 |
| MBP (mm Hg) | 84 ± 9 | 82 ± 8 | 85 ± 8 |
| Steps/day (steps)<sup>1</sup> | 12,619<sup>a,b**</sup> ± 2,613 | 13,227<sup>a**</sup> ± 2,642 | 12,040<sup>b**</sup> ± 2,445 |
| Estimated VO<sub>2peak</sub> (L/min) | 2.322 ± 0.409 | 2.282 ± 0.368 | 2.503 ± 0.547 |
| Estimated VO<sub>2peak</sub> (ml/min/Kg) | 34.29<sup>a*</sup> ± 7.79 | 32.57<sup>b*</sup> ± 7.27 | 32.01<sup>b*</sup> ± 8.18 |
| ΔWG 3T-1T (Kg) |  |  | 10.3 ± 3.7 |
| ΔWG 3T-1T percentage (%) |  |  | 15.4 ± 6.2 |

Values are means and standard deviation; n=30. 1T, first trimester; 2T, second trimester; 3T third trimester; BMI, body mass index ; HR, heart rate; HRR, heart rate reserve; MBP mean blood pressure; <sup>1</sup>, n=26 at 1T, n=25 at T2 and n= 26 at T3 (valid n=23); VO<sub>2peak</sub>, peak oxygen uptake; ΔWG 3T-1T, the weight difference between visit 1 and visit 3; a, b, c letters indicate significant statistical differences between trimesters; \*\*, P <0.001, \*, P < 0.05.

Healthy and unhealthy weight gain groups were similar in age, blood pressure and BMI in the first trimester of pregnancy, but the healthy weight gain group was slightly more active. Conversely, as pregnancy progressed , the unhealthy group had higher BMI, blood pressure and resting HR (Supplemental table 1, Supplemental Digital Content).

Intensity internal load variables of CPE-T during pregnancy are described in Supplemental table 2, Supplemental Digital Content. There was a significant increase in all of them in S2 and S3 throughout pregnancy.

Net WEE and HR increased significantly within speeds and trimesters (figures 2A,2B). Hence, net WEE grew at higher rates between 2^nd^ and 3^rd^ trimester compared to the 1^st^ and 2^nd^ trimester in absolute (table 2) and relative units (S1, 2^nd^-1^st^ = -1.71 ± 2.93% vs. S1, 3^rd^– 2^nd^ = 16.16 ± 3.46%, *P=*0.04; S2, 2^nd^-1^st^= 2.43 ± 2.0% vs. S2, 3^rd^-2^nd^= 16.12 ± 2.61%, *P*<0.001; S3, 2^nd^-1^st^= 6.81 ± 3.47% vs. S3, 3^rd^-2^nd^= 21.79 ± 3.06%, *P=*0.003). In addition, an interaction effect (trimester x speed) was observed for Net WEE (table 2) and RER (figure 2C and Supplemental table 3, Supplemental Digital Content), *F*=22.329*, P<*0.001; *F*=10.949*, P<*0.001, respectively; thus, the increase in Net WEE was higher at S3 than S1 and S2 and RER decreased in S1 and S2 and increased in S3 throughout trimesters.

**Figure 2.**
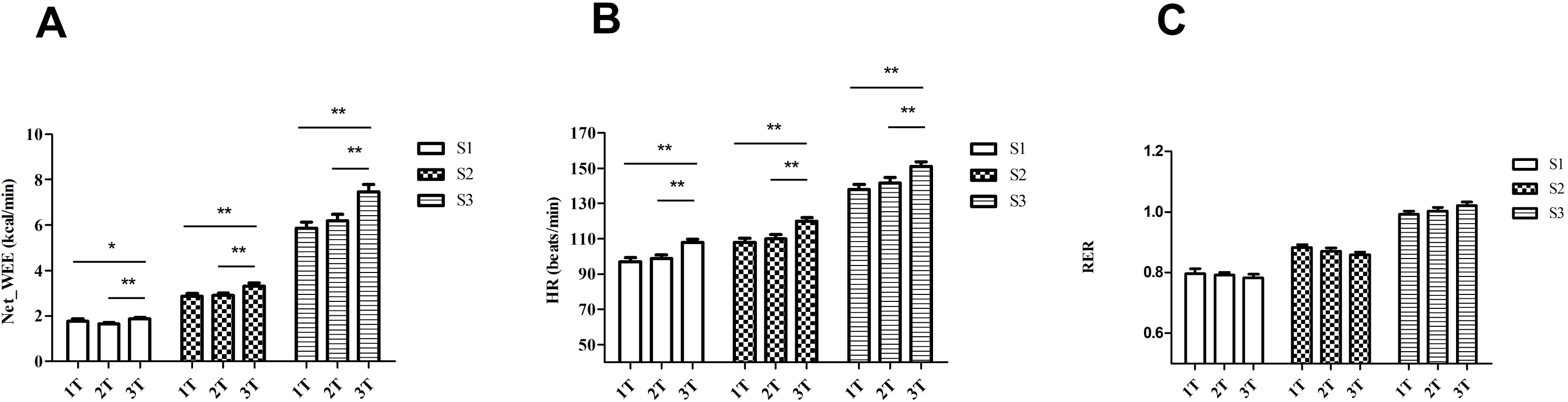
Changes in net walking energy expenditure and heart rate in each speed during pregnancy. Panel A. Changes in net walking energy expenditure at each speed during pregnancy. Net_WEE, net walking energy expenditure in kcal/min; *, P < 0.05; **, P < 0.001; S1, speed 1; S2, speed 2; S3, speed 3; 1T, first trimester; 2T, second trimester; 3T, third trimester. Panel B. Changes in heart rate at each speed during pregnancy. HR, heart rate in beats/min; **, P < 0.001; S1, speed 1; S2, speed 2; S3, speed 3; 1T, first trimester; 2T, second trimester; 3T, third trimester. Panel C. Respiratory exchange ratio. S1, speed 1, S2, speed 2; S3, speed 3; 1T first trimester; 2T second trimester; 3T third trimester.

**Table 2.**
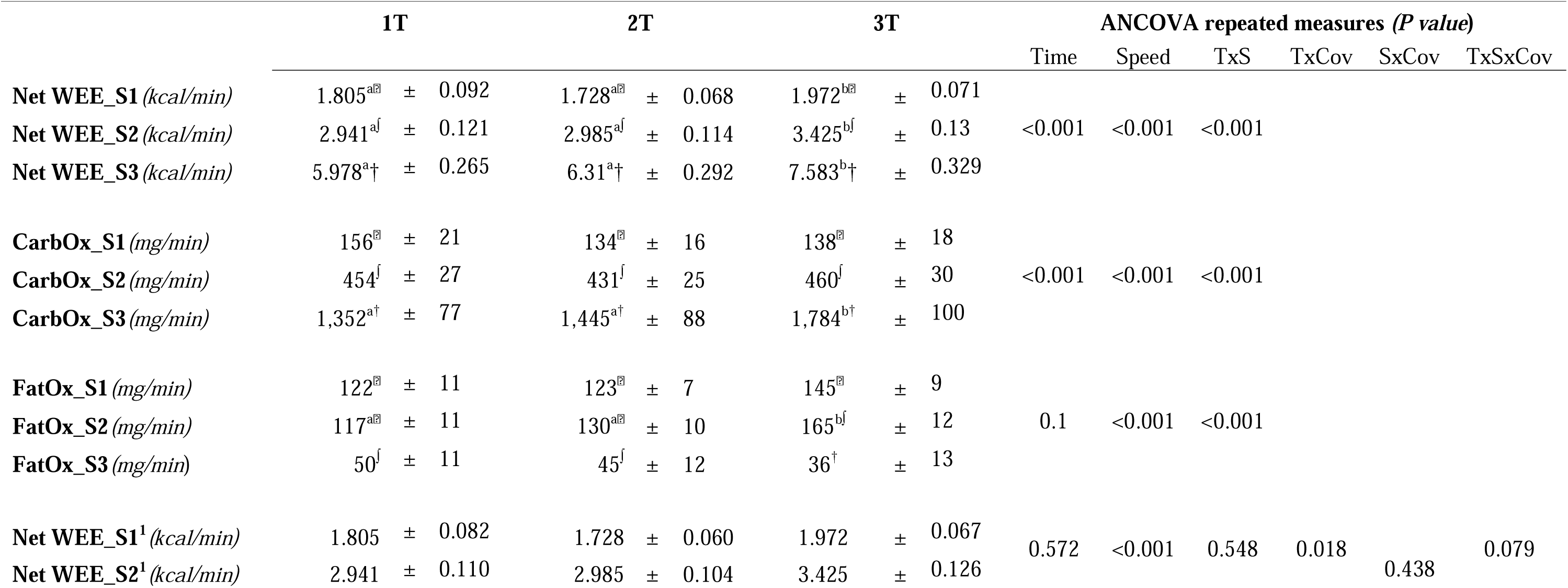
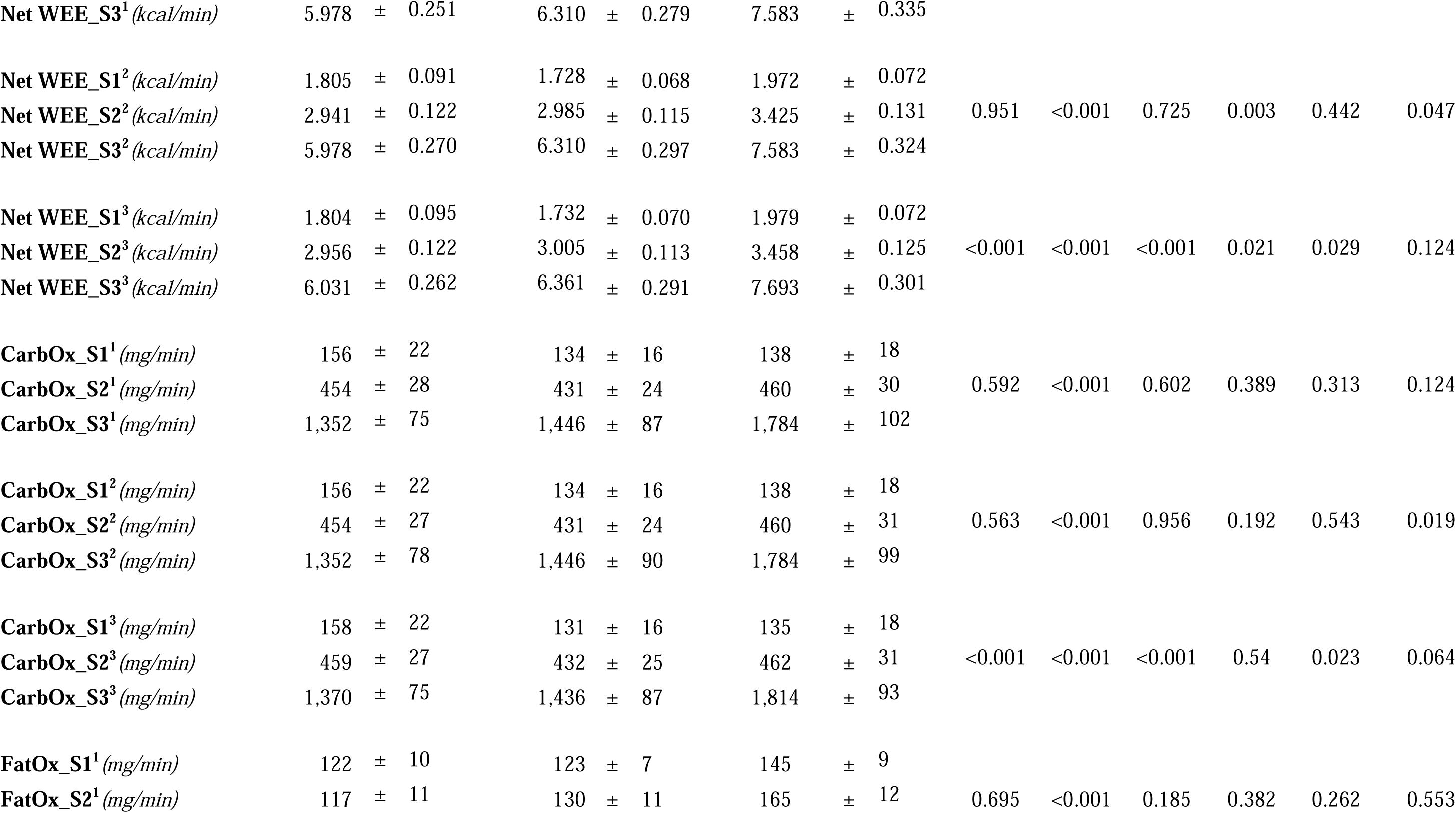
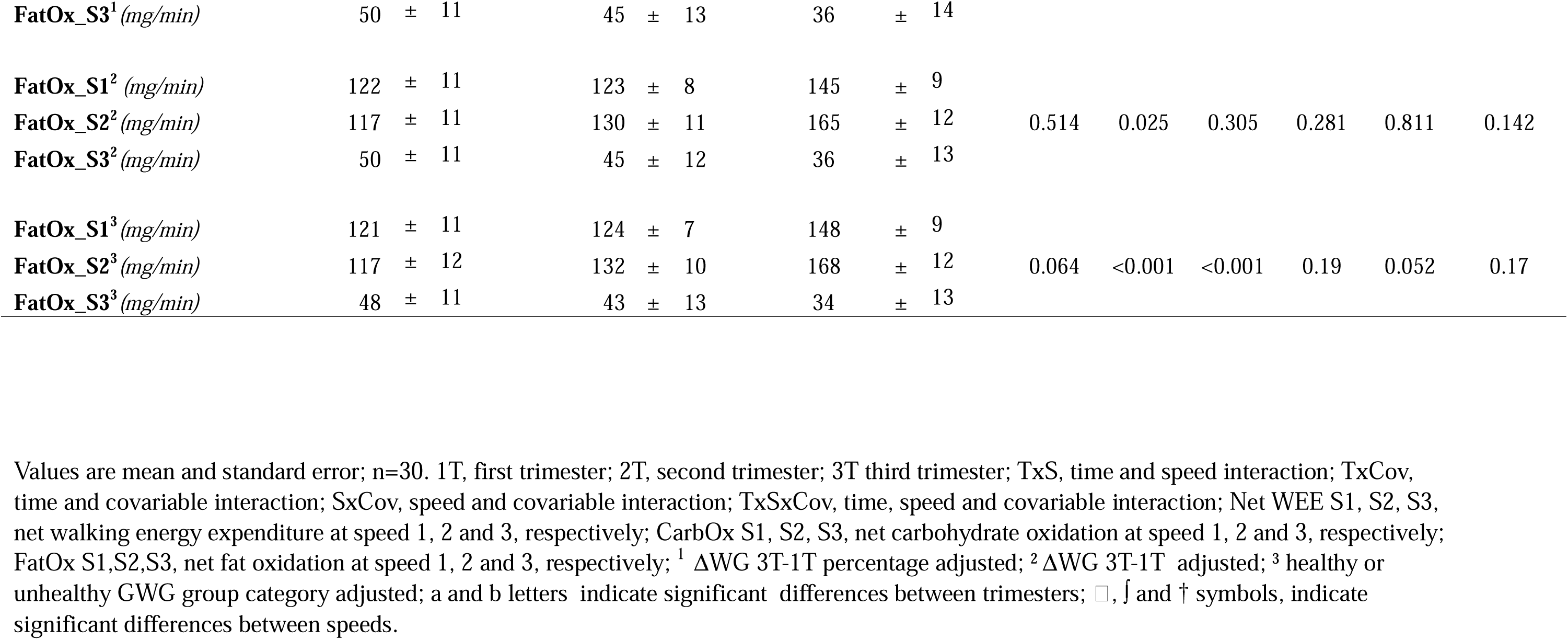
Net walking energy expenditure and substrate oxidation at three walking speeds (S1= 2 Km/h; S2= 4 Km/h; S3= 6 Km/h) during pregnancy.

Fat oxidation (FatOx) and carbohydrate oxidation (CarbOx) rates changed as a function of speed/intensity across trimesters (figure 3). Therefore, at moderate intensity (S2) FatOx increased at the third trimester (3T) compared with the first (1T) and the second one (2T) (figure 3A); conversely, at high walking intensity (S3) only CarbOx increased significantly in 3T compared with the previous trimesters (figure 3B). These results were supported by a significant interaction between trimester and speed when substrates were expressed in absolute (table 2) and relative units (*F*=2.885, *P =* 0.026, figure 3C, respectively). All together, these previous results suggested an intensity-dependent substrate selection during walking exercise throughout pregnancy.

**Figure 3.**
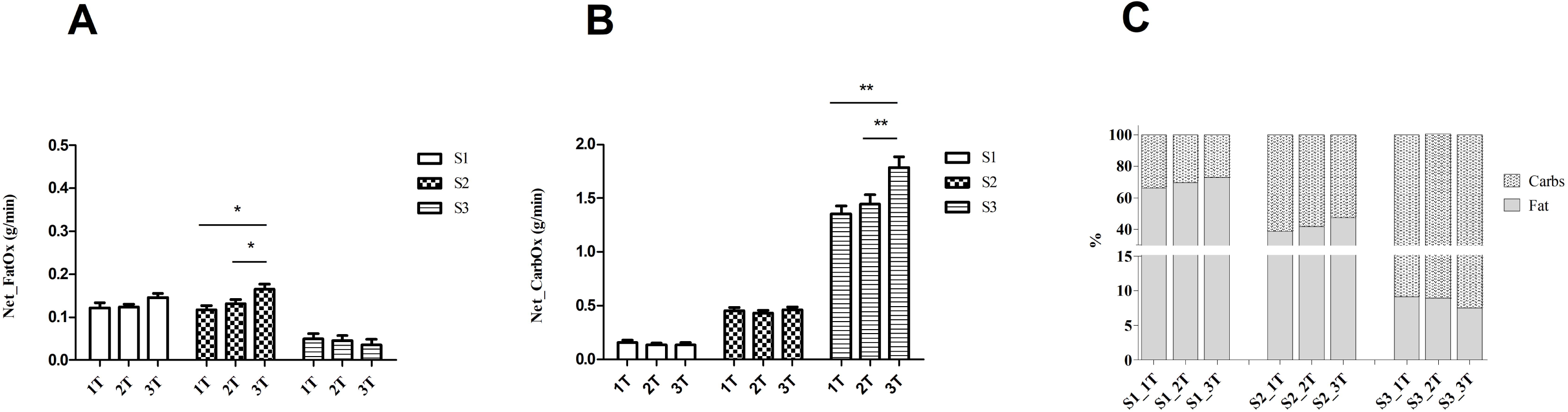
Changes in substrate oxidation at each speed during pregnancy. Panel A. Differences in Fat oxidation; Net_FatOx, net fat oxidation; S1, speed 1; S2, speed 2; S3, speed 3; 1T, first trimester; 2T, second trimester; 3T, third trimester; *, P < 0.05. Panel B. Differences in Carbohydrate oxidation; Net_CarbOx, net carbohydrates oxidation; S1, speed 1; S2, speed 2; S3, speed 3; 1T, first trimester; 2T, second trimester; 3T, third trimester; **, P < 0.001. Panel C. Percentages of substrate use; S1_1T, speed 1 at first trimester; S1_2T, speed 1 at second trimester; S1_3T, speed 1 at third trimester; S2_1T, speed 2 at first trimester; S2_T2, speed 2 at second trimester; S2_T3, speed 2 at third trimester;S3_1T, speed 3 at first trimester;S3_T2, speed 3 at second trimester; S3_T3, speed 3 at third trimester.

All differences between Net WEE and substrate oxidation rates across trimesters were blunted after adjusting by ΔWG 3T-1T or ΔWG 3T-1T percentage (table 2), which suggests quantitative differences in Net WEE and CarbOx were due to ΔWG 3T-1T (*P=*0.047; *F*=3.115 and *P=*0.019; *F*=3.782, for Net WEE and CarbOx respectively). Additionally, analyses by GWG group showed the change in Net WEE was different between groups (table 2) with significant interaction effect between GWG group and speed (*F*= 5.076, *P*= 0.029) and between GWG group and trimester (*F* = 4.162, *P =*0.021). Therefore, the unhealthy weight gain group increased more Net WEE from S1 to S2 and S3 than healthy weight group; also, differences in Net WEE between 3T-1T and 3T-2T were higher in unhealthy than healthy group at S2 and S3 respectively (figure 4).

**Figure 4.**
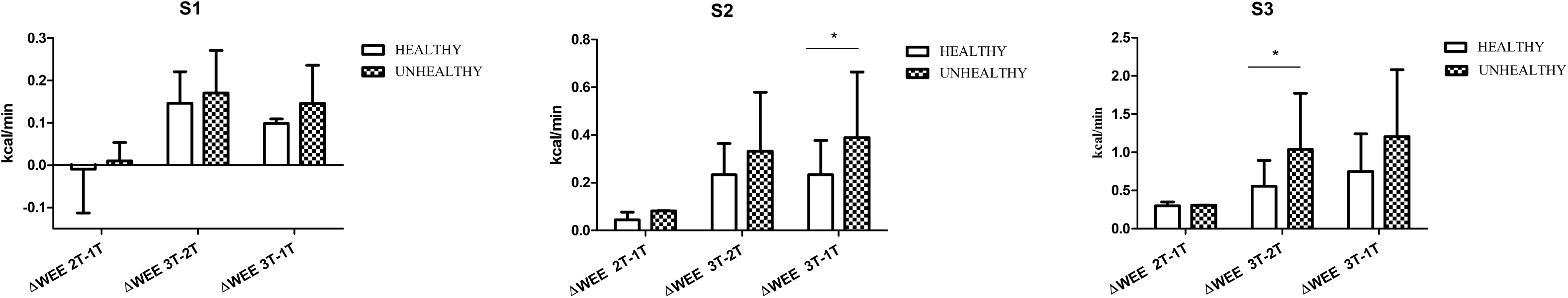
Changes in net walking energy expenditure across trimesters in healthy and unhealthy weight gain groups. Panel A. Changes in speed 1. S1, speed 1; ΔWEE, difference in walking energy expenditure between trimesters; 1T, first trimester; 2T, second trimester; 3T, third trimester. Panel B. Changes in speed 2. S2, speed 2; ΔWEE, difference in walking energy expenditure between trimesters; 1T, first trimester; 2T, second trimester; 3T, third trimester; * P < 0.05. Panel C. Changes in speed 3. S3, speed 3; ΔWEE, difference in walking energy expenditure between trimesters; 1T, first trimester; 2T, second trimester; 3T, third trimester; * P < 0.05.

Regarding substrate oxidation, speed x GWG group interaction for CarbOx, showed the unhealthy group expended significantly more carbohydrates than the healthy group at any given speed (*F*=5.682; *P=*0.023) (table 2).

### Walking Exercise Efficiency

There was a significant interaction trimester x speed for Gross ExE (*P=*0.001) and Net ExE (*P=*0.037) , which suggested walking exercise efficiency changed differently across speeds and trimesters. Thus, the highest efficiency happened always at S2 (moderate intensity) independently of the trimester (table 3). Overall delta efficiency decreased 1.48% from 2T to 3T and 1.88% from 1T to 3T (figure 5).

**Table 3.**
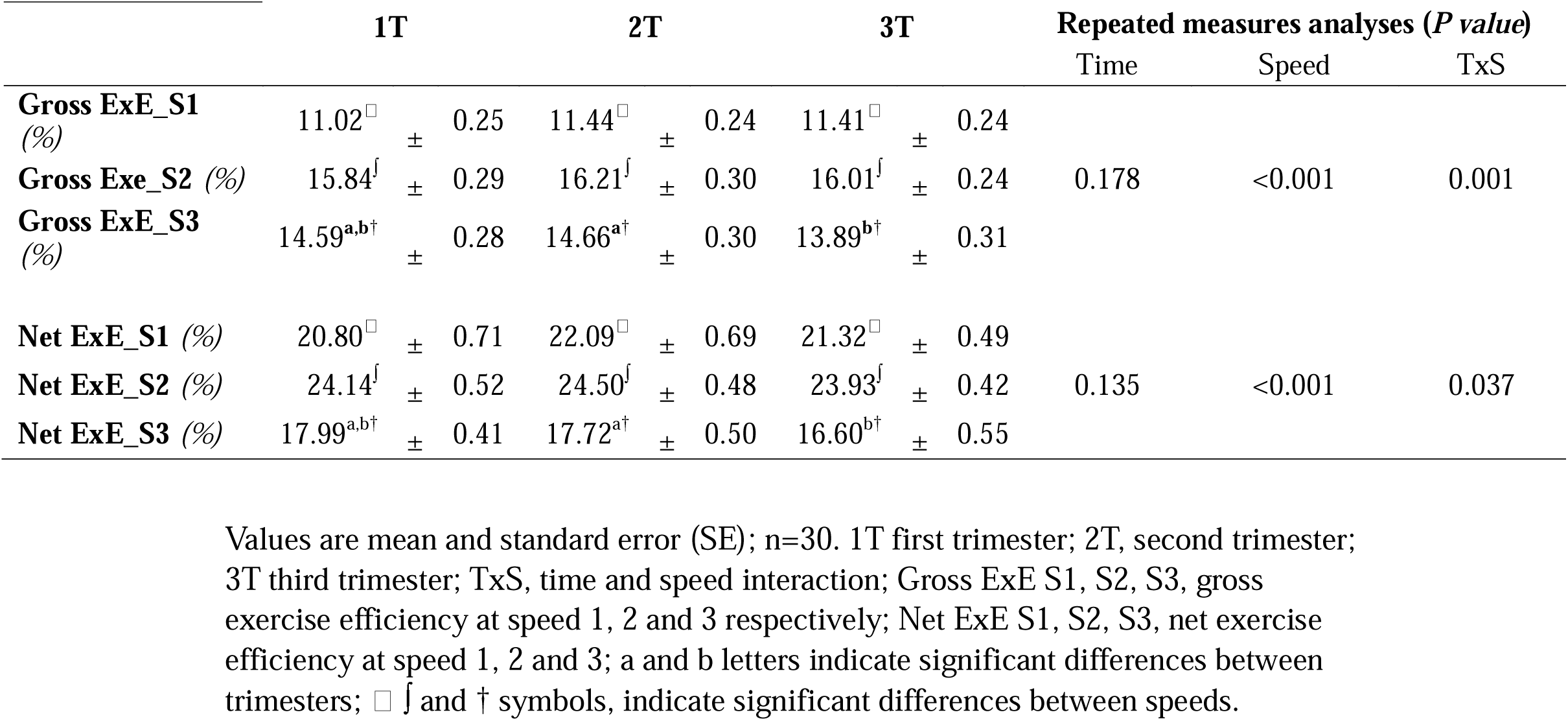
Gross and net exercise efficiency across the three trimesters of pregnancy.

**Figure 5.**
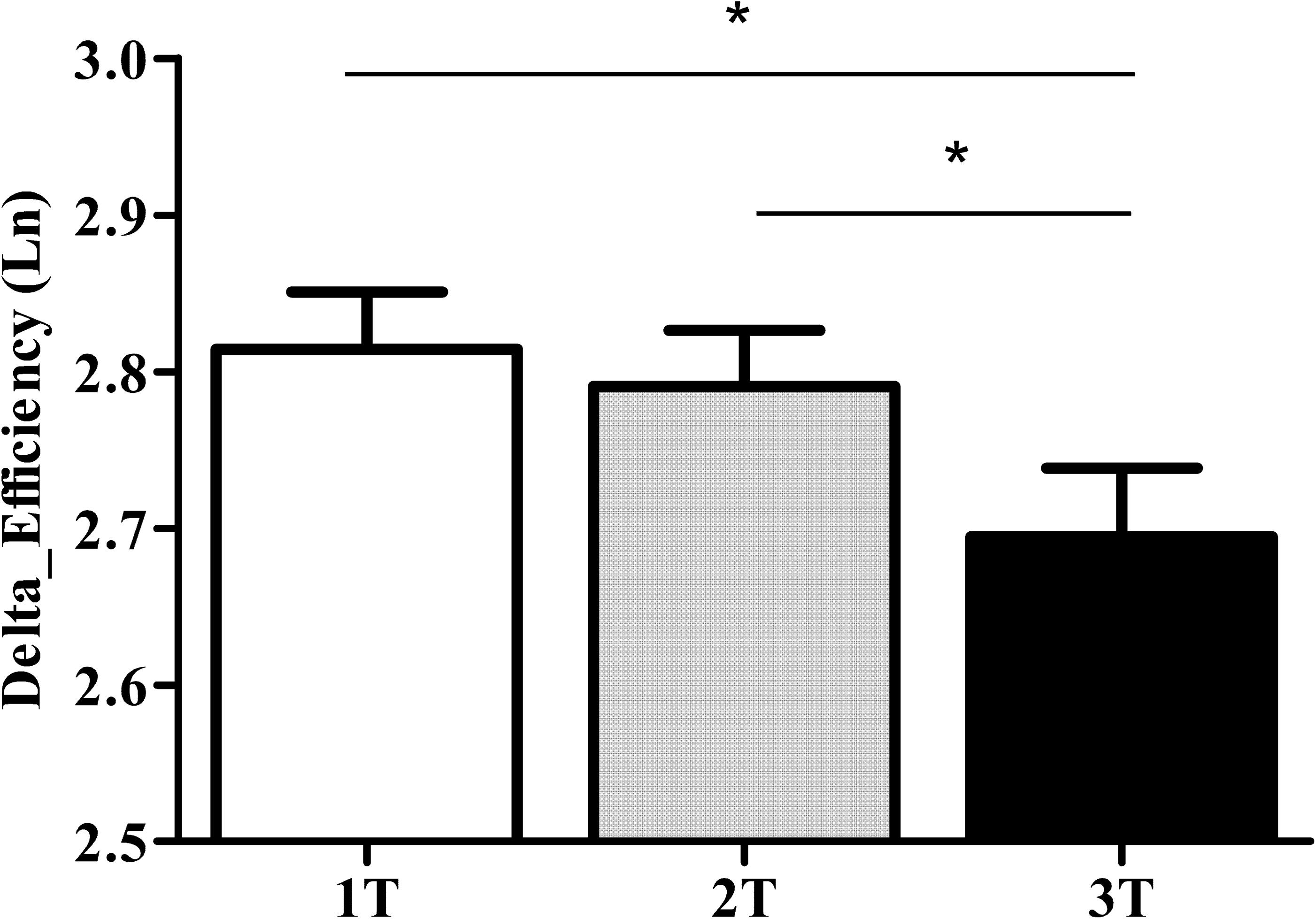
Delta exercise efficiency across the three trimesters of pregnancy. Ln, natural logarithm; 1T, first trimester; 2T, second trimester; 3T, third trimester; *, P < 0.05.

## Discussion

The main finding of this study was a paradoxical change in substrate oxidation selection during exercise during the pregnancy that is intensity dependent. Although, we observed an increase in Net WEE throughout the trimesters at any given speed; pregnant women supported this increase by oxidating more fat only at moderate exercise intensity without significant changes in carbohydrates oxidation. Conversely, at the highest intensity, the increase of energy expenditure was supported by a significant increase in carbohydrates oxidation. Therefore, an increase in relative intensity of exercise during pregnancy is not always supported by an increase in carbs oxidation as we could have inferred from data of non-gestational weight gain (13). This finding may be a specific adaptation of pregnant women which spares the flux of carbohydrates into placental circulation at moderate exercise intensity, suggesting a protecting mechanism of the fetus nervus system fuel (14) when maximum blood flow is not required to support exercise intensity. In fact, in late gestation rising concentrations of somammotropin (HCS), prolactin, cortisol and glucagon exert antiinsulinogenic and lipolytic effects that promote greater use of alternative fuels, specially fatty acids, by peripheral tissues (15).

The changes observed in substrate oxidation were also confirmed by an interaction time x speed in RER ; therefore, RER tended to decrease at light/moderate intensity and to increase at vigorous exercise intensity during pregnancy. These results are in agreement with a previous study that found a significant decrease in RER walking on a treadmill at the same intensity (3.9 km/h, 0% slope), during five longitudinal assessments throughout pregnancy (16). Participants’ characteristics in both studies are similar, although our sample had slightly more BMI and age in early pregnancy.

Increased Net WEE across the three trimesters of pregnancy has been reported previously (16–19). Reductions in Net WEE found in other studies (20) must have been related with methodological differences between the studies; e.g. we used a constant speed while in Byrnés study a self-selected speed was utilized.

The significant elevation in Net WEE in not-trained pregnant women can be related with weight gain, impaired in exercise efficiency and other physiological adaptations related with pregnancy (14,17,18,21); in agreement with these previous studies, we confirm the relationship between weight gain and Net WEE.

Regarding efficiency, we found a significant decrease in delta efficiency during pregnancy. Reductions in efficiency have been related with switching from fat to carbohydrate substrate oxidation at the same absolute exercise load (22), which must be related to an impaired exercise metabolic flexibility. A deeper analysis of exercise efficiency data showed only a significant decrease in gross and net efficiency at the highest intensity of walking at the third trimester compared with the second one, which coincided with an increase in carbohydrate oxidation at the highest speed. These results highlight the relevance of exercise efficiency as a marker of metabolic health described in non-pregnant populations (23). Comparing with pregestational values, lower (17) and higher efficiency (24) was found during pregnancy, especially in well conditioning pregnant women (24).

Weight gain is an important variable in net WEE and substrate oxidation throughout pregnancy. We found an interaction time x speed x ΔWG 3T-1T that could explain changes in Net WEE and CarbOx. Other studies showed that pregnant women with higher BMI had more energy expenditure cost during pregnancy (25) or a strong positive correlation between weight gain during pregnancy and the change in energy expenditure per activity (17). Even in no pregnant population some studies showed that energy expenditure increases when participants lift a load during walking (26,27) or even standing (27) .Weight gain can also affects walking biomechanics; thus, the expected increase in BMI has been suggested as a primary factor linking alteration on walking mechanics and increased walking energy expenditure (21).

As we noted above, despite the weight gain during pregnancy, FatOx increased at moderate intensity of walking during pregnancy, as a more efficient way to support exercise energy cost. Hence, the highest efficiency was found walking at 4 km/h in all trimesters. This finding is totally different to what we expect in non-gestational populations as skeletal muscle work efficiency decreases after weight gain (13).

To analyze the effect of being into IOM recommendations in WEE and substrate oxidation, participants were classified in a healthy or unhealthy gestational weight group. As we hypothesized, the unhealthy group showed less efficiency than the healthy group, so they spent more Net WEE throughout pregnancy than the healthy group, especially walking at 4 and 6 km/h. Also, the unhealthy group used more carbs at any given speed.

## Strengths and limitations

The main strength of this study was that we perform longitudinal assessments of exercise energy expenditure and substrate oxidation during pregnancy in the same group of women. Moreover, we provided metrics of exercise net energy expenditure and efficiency to remove the well-known increase in metabolic rate associated with pregnancy (17,20,25,28) , which can confound exercise energy expenditure output (29). However, we acknowledge that a standard assessment of resting metabolic rate (RMR) could be performed to calculate Net WEE.

The results of this study can be applied to white women and different conclusions might have been obtained with participants from other ethnic backgrounds.

Since we had an estimation of VO_2peak_, we were not able to analyze the impact of VO_2peak_ in the differences on WEE and substrate oxidation during pregnancy. Therefore, the influence of cardiorespiratory fitness and physical activity on changes in WEE and substrate oxidation during pregnancy should be addressed in future studies.

## Conclusion

In summary, our analyses described a paradoxical reversal switching in substrate oxidation during exercise during pregnancy which is intensity dependent. Therefore, the increase in walking energy expenditure after the second trimester (at the same speed) relies on FatOx without changes in CarbOx. This adaptation translates in impaired exercise efficiency after the second trimester at high walking intensities compared to moderate-low intensities. These changes seem to be more pronounced in women with excessive weight gain.

These results may be relevant for the management and treatment of gestational obesity, weight gain, and diabetes during pregnancy, which are cornerstone strategies to improve maternal and neonatal health outcomes.

## Data Availability

All data produced in the present study are available upon reasonable request to the authors

