## Supplemental Figure 1 for "Changes in Walking Energy Expenditure and Substrate Oxidation during Pregnancy"

Figure 1. Flowchart

47 pregnant women agree to participate

First trimester

39 valid measures

Second trimester

36 valid measures

Third trimester

30 valid measures

50 pregnant women recruited

3 losses:

2 miscarriages

1 give up

3abandened

6 losses:

1 preterm delivery

1 miscarriage

2 configuration fails

2 give up

30 pregnant women with 3 valid measures

8 losses:

4 miscarriages

1 configuration fail

3 give up
