## Supplemental Table 1 for "Changes in Walking Energy Expenditure and Substrate Oxidation during Pregnancy"

Table 1. Healthy and unhealthy gestational weight gain groups descriptive characteristics (n= 17 in healthy group, n=13 in unhealthy group)

|  |  | 1T | |  | 2T | |  | 3T | |
| --- | --- | --- | --- | --- | --- | --- | --- | --- | --- |
|  |  | Healthy | Unhealthy |  | Healthy | Unhealthy |  | Healthy | Unhealthy |
|  |  | Mean SD | Mean SD |  | Mean SD | Mean SD |  | Mean SD | Mean SD |
| Age *(years)* | | 33.4 ± 4.7 | 33.1 ± 5.4 |  |  |  |  |  |  |
| Gestational week *(weeks)* | | 10.7± 1.1 | 10.2 ± 1.9 |  | 17.06 ± 0.9 | 18.08 ± 1.8 |  | 32.7 ± 1.6 | 33.1 ± 1.5 |
| BMI *(Kg/m²)* | | 25.0 ± 4.3 | 28.4 ± 5.8 |  | 25.6^a*^ ± 3.9 | 29.9^b*^± 5.9 |  | 27.96^a*^ ± 3.77 | 33.5^b*^ ± 5.41 |
| HR resting *(beats/min)* | | 78.9 ± 15.2 | 83.6 ± 11.2 |  | 78.1^a*^ ± 11.7 | 85.3^b*^± 9.1 |  | 88.8^a*^ ± 10.1 | 95.4^b*^ ± 9.9 |
| HRR *(beats/min)* | | 105.7 ± 16.0 | 101.3 ± 8.6 |  | 106.6^a*^± 12.8 | 99.6^b*^ ± 8.0 |  | 95.5^a*^ ± 9.7 | 89.4^b*^ ± 9.5 |
| MBP *(mm Hg)* | | 83.9 ± 9.49 | 84.2 ± 7.37 |  | 80.2 ± 7.49 | 83.9 ± 7.29 |  | 81.8^a*^ ± 7.13 | 88^b*^ ± 6.72 |
| Steps/day *(steps)¹* | | 13529^a*^± 2375 | 11558^b*^± 2563 |  | 13456 ± 2348 | 12934 ± 3069 |  | 12469 ± 2507 | 11454 ± 2344 |
| Estimated VO2peak *(L/min)* | | 2.356 ± 0.398 | 2.279 ± 0.436 |  | 2.323 ± 0.382 | 2.229 ± 0.356 |  | 2.545 ± 0.526 | 2.448 ± 0.589 |
| ΔWG 3T-1T *(Kg)* | |  |  |  |  |  |  | 10.5^a**^ ± 2.7 | 15.8^b**^ ± 3.9 |
| ΔWG 3T-1T percentage *(%)* | |  |  |  |  |  |  | 13.2^a*^ ± 5.1 | 18.4^b*^ ± 6.3 |

Values are means and standard deviation; 1T, first trimester; 2T, second trimester; 3T third trimester; BMI, body mass index ; HR, heart rate; HRR, heart rate reserve; MBP mean blood pressure; ¹, n=14 and 12 in healthy and unhealthy group respectively in 1T, n=14 and 11 in healthy and unhealthy group respectively in 2T , n=15 and 11 in healthy and unhealthy group respectively in 3T; VO_2 peak_, peak oxygen uptake; ΔWG 3T-1T, the weight difference between visit 1 and visit 3; a, b letters indicate significant statistical differences between groups; **, P <0.001, *, P < 0.05.
