## Supplemental Table 2 for "Changes in Walking Energy Expenditure and Substrate Oxidation during Pregnancy"

Table 2. Intensity load variables of CPE-T during pregnancy

|  | 1T | | | 2T | | | 3T | | |
| --- | --- | --- | --- | --- | --- | --- | --- | --- | --- |
| %VO2peak S1 | 29.26 | ± | 6.52 | 29.55 | ± | 6.18 | 30.44 | ± | 6.64 |
| %VO2peak S2 | 39.38 | ± | 7.34 | 40.63 | ± | 7.67 | 42.45 | ± | 9.04 |
| %VO2peak S3 | 61.78^a^ | ± | 12.17 | 64.49^a,b^ | ± | 13.27 | 68.33^b^ | ± | 15.08 |
|  |  | ± |  |  | ± |  |  | ± |  |
| %VO2R S1 | 18.40 | ± | 5.42 | 18.42 | ± | 5.08 | 19.46 | ± | 5.18 |
| %VO2R S2 | 30.11^a^ | ± | 6.73 | 31.32^a,b^ | ± | 7.04 | 33.48^b^ | ± | 8.47 |
| %VO2R S3 | 56.12^a^ | ± | 13.03 | 59.16^a,b^ | ± | 14.33 | 63.71^b^ | ± | 16.32 |
|  |  | ± |  |  | ± |  |  | ± |  |
| %HRmax S1 | 51.94^a^ | ± | 3.92 | 53.02^a^ | ± | 1.13 | 58.15^b^ | ± | 1.10 |
| %HRmax S2 | 57.89^a^ | ± | 1.14 | 59.06^a^ | ± | 1.13 | 64.63^b^ | ± | 1.10 |
| %HRmax S3 | 74.14^a^ | ± | 1.13 | 75.98^a^ | ± | 1.14 | 81.26^b^ | ± | 1.11 |
|  |  | ± |  |  | ± |  |  | ± |  |
| %HRR S1 | 13.62 | ± | 1.78 | 15.98 | ± | 1.42 | 16.78 | ± | 1.49 |
| %HRR S2 | 25.08^a^ | ± | 1.34 | 26.87^a,b^ | ± | 1.33 | 29.46^b^ | ± | 1.36 |
| %HRR S3 | 53.79^a^ | ± | 1.27 | 57.58^b^ | ± | 1.29 | 62.12^c^ | ± | 1.30 |
| RPE (Borg scale) S1 | 1 |  |  | 1 |  |  | 1 |  |  |
| RPE (Borg scale) S2 | 2 |  |  | 2 |  |  | 3 |  |  |
| RPE (Borg scale) S3 | 4 |  |  | 4 |  |  | 5 |  |  |

Values are mean and standard deviation except in Borg scale, where data are in median; n=30. 1T, first trimester; 2T, second trimester; 3T, third trimester; %VO_2peak_, peak oxygen consumption percentage; %VO_2_R, oxygen consumption reserve percentage; %HR_max_, maximal heart rate percentage; %HRR heart rate reserve percentage; RPE, rating of perceived exertion; S1, S2, S3 speed 1, 2 and 3; a,b,c letters indicate significant differences between trimesters (*P*<0.05).
