## Supplemental Table 3 for "Changes in Walking Energy Expenditure and Substrate Oxidation during Pregnancy"

Table 3. Changes in RER during pregnancy

|  | 1T | | |  | 2T | | |  | 3T | | | Repeated measures analyses (*P value*) | | |
| --- | --- | --- | --- | --- | --- | --- | --- | --- | --- | --- | --- | --- | --- | --- |
|  |  |  | |  |  |  | |  |  |  | | Time | Speed | TxS |
| RER_S1 | 0.835^Ɵ^ | ± | 0.01 |  | 0.828^Ɵ^ | ± | 0.01 |  | 0.825^Ɵ^ | ± | 0.01 | 0.908 | <0.001 | <0.001 |
| RER_S2 | 0.881^∫^ | ± | 0.01 |  | 0.871^∫^ | ± | 0.01 |  | 0.864^∫^ | ± | 0.01 |  |  |  |
| RER_S3 | 0.967^†^ | ± | 0.01 |  | 0.973^†^ | ± | 0.01 |  | 0.991^†^ | ± | 0.01 |  |  |  |

Values are mean and standard error (SE); n=30. 1T first trimester; 2T, second trimester; 3T third trimester; TxS, time and speed interaction; RER_S1, Respiratory exchange ratio in speed 1; RER_S2, Respiratory exchange ratio in speed 2; RER_S3, Respiratory exchange ratio in speed 3; Ɵ ∫ and † symbols, indicate significant differences between speeds.
